# Association of dietary patterns and body phenotypes in Brazilian adolescents

**DOI:** 10.1101/2021.05.14.21257150

**Authors:** Ana Elisa Madalena Rinaldi, Wolney Lisboa Conde, Carla Cristina Enes

## Abstract

**Objective:** To investigate the association between dietary patterns and physical activity and body phenotypes in Brazilian adolescents.

**Methods:** Cross-sectional study using data from a school-based study with 1,022 Brazilian adolescents aged 10 to 19 years. Body phenotypes (BP) and dietary patterns (DP) were defined using principal components analysis. BP was defined using anthropometric, body composition, biochemical, and sexual maturation, and DP from 19 food groups categorized from a food frequency questionnaire containing 58 food items. We performed a scatter plot to describe the relationship between energy expenditure assessed by MET and BP scores. The association between DP and BP, adjusted by sex, age and socioeconomic status, was assessed by linear regression model.

**Results:** We identified five BP (BP1_adiposity_, BP2_puberty_, BP3_biochemical_, BP4_muscular_, BP5_lipids_biochemical_) and five DP (DP1_ultraprocessed_foods,_ DP2_fresh_foods_, DP3_bread_rice_beans_, DP4_culinary_preparation,_ DP5_cakes_rice_beans_). There are higher scores of BP_adiposity for obese adolescents, but energy expenditure was similar for obese and non-obese. Physical activity was positively associated with body mass index, BP_adiposity and BP_puberty. We observed negative association between DP_ultraprocessed and BMI, and a positive between DP_fresh_foods. DP_fresh_foods was positively associated with BP_adiposity; DP_ultraprocessed and DP_culinary_preparation were negatively associated with this BP. BP_biochemical was negatively associated with DP_fresh_foods.

**Conclusion:** We identified negative association between DP mainly composed by ultraprocessed foods and fresh foods and BP_adiposity._ These associations need to be more explored, especially in adolescents, because both DP and BP were defined using multivariate analysis.

## Introduction

Healthy eating practices during adolescence have a profound impact on the immediate and long-term future health of adolescents and may limit harmful behaviors contributing to the epidemic of non-communicable diseases (NCDs) in adulthood.^1,2^ In 2016, considering that nutrition is a central health, economic, and sustainable development challenge, the United Nations implemented the Decade of Action on Nutrition (2016-2025), which aimed to mobilize all governments to accelerate actions to eradicate all forms of malnutrition (undernourishment, micronutrient deficiencies, and overweight / obesity).^3^ In addition, these actions must be coordinated with sustainable development goals and the food system needs to offer a healthy diet for humans and the environment.^4^

Although several evidence-informed actions exist to address nutrition actions relevant to adolescents, there are gaps in understanding their health situation and further research is merited. There are relevant aspects related to adolescent health such as nutrition, physical activity, mental and reproductive health, alcohol, tobacco, and drug use.^5^

Another aspect that can impede a broader understanding of adolescent health is the frequent use of traditional analyses that investigate the isolated effect of food consumption, physical activity and body composition instead of a joint and dynamic assessment of behavioral profiles, dietary patterns (DPs) and the main influencers of adolescents in social and psychosocial contexts.

The nutritional status of adolescents also has an important impact on their health and is usually assessed using isolated indicators – anthropometry, body composition or biochemical markers – that are analyzed separately with social and psychosocial predictors, hindering a broader understanding of the health situation of adolescents.^6,7^ These indicators are unable to address the complexity of body composition changes during puberty. An alternative to this limitation is the use of multivariate analysis for the definition of body phenotypes (BPs) that are the sum of the specificities that characterize an individual.^8^

The use of BPs in the assessment of nutritional status in adolescents is an innovative approach demonstrated by the simultaneous analysis of anthropometric dimensions, body composition and biochemical parameters in a multivariate model. Previous studies^9,10^ have proposed and described BPs using anthropometric, sexual maturation, body composition, and biochemical variables in multivariate analyses. In our study we applied these BPs as outcomes to understand their association with dietary patterns, physical activity, and sociodemographic characteristics. Therefore, the aim of our study was to investigate the association between DPs and physical activity, and BPs in adolescents.

## Methods

### Study population and study design

We performed a cross-sectional study using data from a school-based study entitled “Determinants of the risk of obesity among adolescents from a survey of schoolchildren with a mixed sample: transversal and longitudinal” (IAP-SP). The IAP-SP corresponds to a third wave of surveys conducted with school adolescents in the city of Piracicaba, São Paulo state, Brazil. This is a probabilistic sample of schools, using two stratification criteria: geographic (central and peripheral districts) and school administration type (public and private). Subsequently, the sample was stratified by school grade and the clusters were analyzed in two stages. In the first stage, the primary sampling units were all the public and private schools; and in the second stage, the secondary sampling units were all the grades in each school. The study enrolled 1,022 adolescents aged 10 to 19 years. The study was approved by the Ethics Committee of the University of Sao Paulo (protocol number 02546612.5.0000.5421) and all subjects’ parents gave written informed consent for their child’s participation in the study.

### Anthropometric data, sexual maturation, and biochemical variables

BPs were defined using anthropometric (weight, height, skinfolds, waist circumference), body composition (phase angle), biochemical (total cholesterol, triacylglycerol, glucose, and hemoglobin), sexual maturation (pubic hair, genitals and breasts) variables, and age (years). The same variables have been used in previous studies.^9,10^ The body mass index (BMI) was defined as ratio of the weight of a person to the square of the height.

All anthropometric measurements were performed in duplicate by trained researchers using standard techniques, considering the mean between values.^11,12^ The phase angle was calculated as the arc tangent of the ratio of reactance to resistance. Resistance and reactance were measured with a bioimpedance analyzer (BIA) (model 0358T, RJL Systems, Clinton Township, USA).^13,14^ All procedures were performed according to recommendations in the manufacturer’s manual. All variables were continuous.

The sexual maturation assessment was based on breast development and pubic hair growth for girls, and genital development and pubic hair growth for boys.^15,16^ All adolescents classified themselves in one of the five maturation stages according to line drawings representing the five different Tanner stages. Self-classification has been reported to present similar results to direct evaluation by health professionals in comparative studies.^17^ Adolescents aged 15 years or more did not complete the assessment of sexual maturation. Furthermore, when the girls were asked about age at menarche, we understood that they had completed the sexual maturation process. Therefore, we classified all girls aged 15 years at Tanner stage 5. For the boys, 16% were aged 16 years old and over, and they were also considered to be at Tanner stage 5.

Blood samples were collected in the morning, after 12 hours of fasting, according to blood collection techniques in accordance with the Ministry of Health protocol.^18^ Triglycerides, glucose (fasting glycemia), hemoglobin, total cholesterol, and lipoprotein fractions values were determined in plasma. The total cholesterol / LDL cholesterol ratio was used.

### Dietary assessment and food groups

The usual food intake assessment was performed by applying the validated Semi-Quantitative Food Frequency Questionnaire (SQFFQ) for adolescents.^19,20^ This tool provides seven consumption choices for 58 food items over the past 3 months. Trained fieldworkers conducted face-to-face structured interviews and filled out the questionnaires in the schools. We classified the 58 original food items into 19 groups based on the similarity of nutrient profiles or culinary usage among foods, and some individual food items were classified individually if their composition differed substantially from that of other foods; the resulting classification is as follows: fruits; vegetables; meat/poultry/fish/eggs; processed meat/sausage; snacks; fast foods; breads; fried tubers; dairy foods; fatty foods; sugary drinks; sugar/pastries; ice cream; cookies; bakery foods; cakes; side dishes; rice/beans; and potatoes/corn (Table 1).

**Table 1.** Description of food groups defined from semiquantitative food frequency questionnaire

| **Food groups** | **Original foods** |
| --- | --- |
| Fruits | Apple; mango/papaya; orange/tangerine; strawberry/pineapple; banana; fruit juices |
| Vegetables | Tomato; pumpkin/carrot; broccoli; lettuce; beet; chayote |
| Meat/poultry/fish/eggs | Fried meat, pork, chicken and fish; grilled meat, pork, chicken and fish; fried eggs/scrambled eggs/omelet |
| Processed meat/sausage | Sausages |
| Snacks | Snacks |
| Fast foods | Hot dog; burger, sandwiches |
| Breads | White bread; loaf bread |
| Fried tubers | French fries/fried manioc |
| Dairy foods | Yoghurts; cheese; whole milk; creamy cheese |
| Fatty foods | Margarine; butter; olive oil/oil |
| Sugary drinks | Soft drinks; carbonated drinks; nectar juices |
| Sugar/pastries | Sugar; chocolate powder; goodies; jam/fruits in syrup |
| Ice-cream | Ice-cream; popsicle |
| Cookies | Filled cookies; non-filled cookies |
| Bakery foods | Baked, pizza, |
| Cakes | Cakes; cake mixes |
| Side dishes | Potato salad with mayonnaise; pasta with tomato sauce; pasta with tomato sauce and meat; risoto with chicken or fish |
| Rice/beans | Rice and beans (Brazilian dish) |
| Potatoes/corn | Boiled potatoes; boiled corn |

### Physical activity and sociodemographic variables

Sociodemographic variables included age, sex, ethnicity, school administration (public and private), and socioeconomic status. The score of household assets were applied as a proxy of wealth. A list of household assets and the schooling of the household head was compiled using the Brazilian Economic Classification Criteria.^21^ These variables were modelled using principal component analysis (PCA),^22^ and only the first component was selected to summarize the data, which was partitioned into tertiles. We also assessed usual physical activity (during the last 12 months) using a validated questionnaire for adolescents.^23^ The intensity of physical activity was expressed in metabolic equivalents (METs) using data on the type of physical activity, duration (hours), and weight (in kilograms). We classified METs according to the World Health Organization in three intensity categories, which were as follows: low intensity (1 to 1.9 METs), moderate intensity (3 to 5.9 METs), and vigorous intensity (more than 6 METs).^24^ METs were also used as continuous variable in the logarithmic scale.

### Statistical analysis

BPs were defined by PCA based on anthropometrics, body composition, biochemical variables, and sexual maturation. Components with an eigenvalue ≥ 1.0 were retained and factor loadings > 0.2 were used to describe the phenotypes. The Kaiser-Meyer-Olkin (KMO) test was used to assess sampling adequacy in relation to the degree of correlation among variables. Further details on the construction of BPs have been presented elsewhere.^9^

Dietary patterns (DPs) were defined from 19 food groups using PCA. Components with eigenvalues higher than 1.0 were retained, and eigenvectors (factor loadings) higher than 0.2 were used to describe the DP. The Kaiser-Meyer-Olkin (KMO) test was used to analyze the compliance of variables with the PCA. Each adolescent received a standardized score for each DP identified. The value of the score represents the proximity of the adolescent to each DP.

We performed a scatter plot to describe the relationship between energy expenditure according to MET and BP scores. The association between BPs and DPs, adjusted by sex, age, and socioeconomic status, was assessed using a linear regression model. All analyses were carried out using Stata SE 13.0 software.

## Results

A total of 1,022 adolescents were assessed in this study. The main differences between adolescents from private and public schools were ethnicity and wealth. The percentage of girls was higher than that of boys (55.9% versus 44.1%), and most of the adolescents were under the age of 15 years from public (65.2%) and private schools (66.1%). The percentage of adolescents classified as white was higher in private schools (83.3%), and mixed race (28.9%) and black (17.6%) in the public ones. Most adolescents in public school were classified in the lower tertiles of wealth (44.8%), and in the private ones were the higher ones (80.4%). The percentage of excess weight (overweight and obesity) and low physical inactivity (MET) was 37.9% and 50.0%, respectively, and it was similar in the public and private schools (data not shown in tables).

Five BPs were defined: BP1_adiposity_, characterized by positive loadings for the skinfold, body mass, and waist circumference variables; BP2_puberty_, characterized by positive loadings for stages of pubic hair and breasts in girls, or pubic hair and genitals in boys, height, and age; BP3_biochemical_, characterized by positive loadings for triglycerides and glucose; BP4_muscule_, characterized by positive loadings for phase angle and hemoglobin; and BP5_lipids_biochemical_ with negative loadings for phase angle and positive ones for triglycerides and the total cholesterol / LDL cholesterol ratio. The KMO value was high (0.8096) (Table 2).

**Table 2.** Body phenotypes (BP) defined from demographic, anthropometric, body composition and biochemical variables for adolescents. Piracicaba, São Paulo, Brazil, 2012.

| Variables | Eigenvalues |  |  |  |  |
| --- | --- | --- | --- | --- | --- |
|  | BP_adiposity | BP_puberty | BP_biochemical | BP_muscular | BP_lipids_biochemical |
| Weight (kg) | 0.4313 |  |  |  |  |
| Height (cm) |  | 0.3840 |  |  |  |
| Tricipital skinfold (mm) | 0.4897 |  |  |  |  |
| Subscapular skinfold (mm) | 0.5165 |  |  |  |  |
| Waist circumference (cm) | 0.5017 |  |  |  |  |
| Phase angle (°) |  |  |  | 0.2538 | -0.3623 |
| Pubic hair (sexual maturation) |  | 0.5099 |  |  |  |
| Gonads and breast (sexual maturation) |  | 0.5126 |  |  |  |
| Hemoglobin (mg/dL) |  |  |  | 0.9204 |  |
| Triglycerides (mg/dL) |  |  | 0.2360 |  | 0.2932 |
| Glucose (mg/dL) |  |  | 0.9270 |  |  |
| Total cholesterol/LDL-chol |  |  |  |  | 0.8664 |
| Age (years) |  | 0.5079 |  |  |  |
| **KMO** |  |  | **0.8096** |  |  |

The relationship between the intensity of physical activity (METs) and BP according to sex and obesity diagnosis is described in Figure 1. For all BPs, it was possible to define two groups of adolescents – physically inactive (energy expenditure < 1.0) and physically active (energy expenditure ≥ 3.0). Higher scores of BP__adiposity_ are presented for obese adolescents, but energy expenditure (log) was similar for both obese and non-obese adolescents.

**Figure 1.**
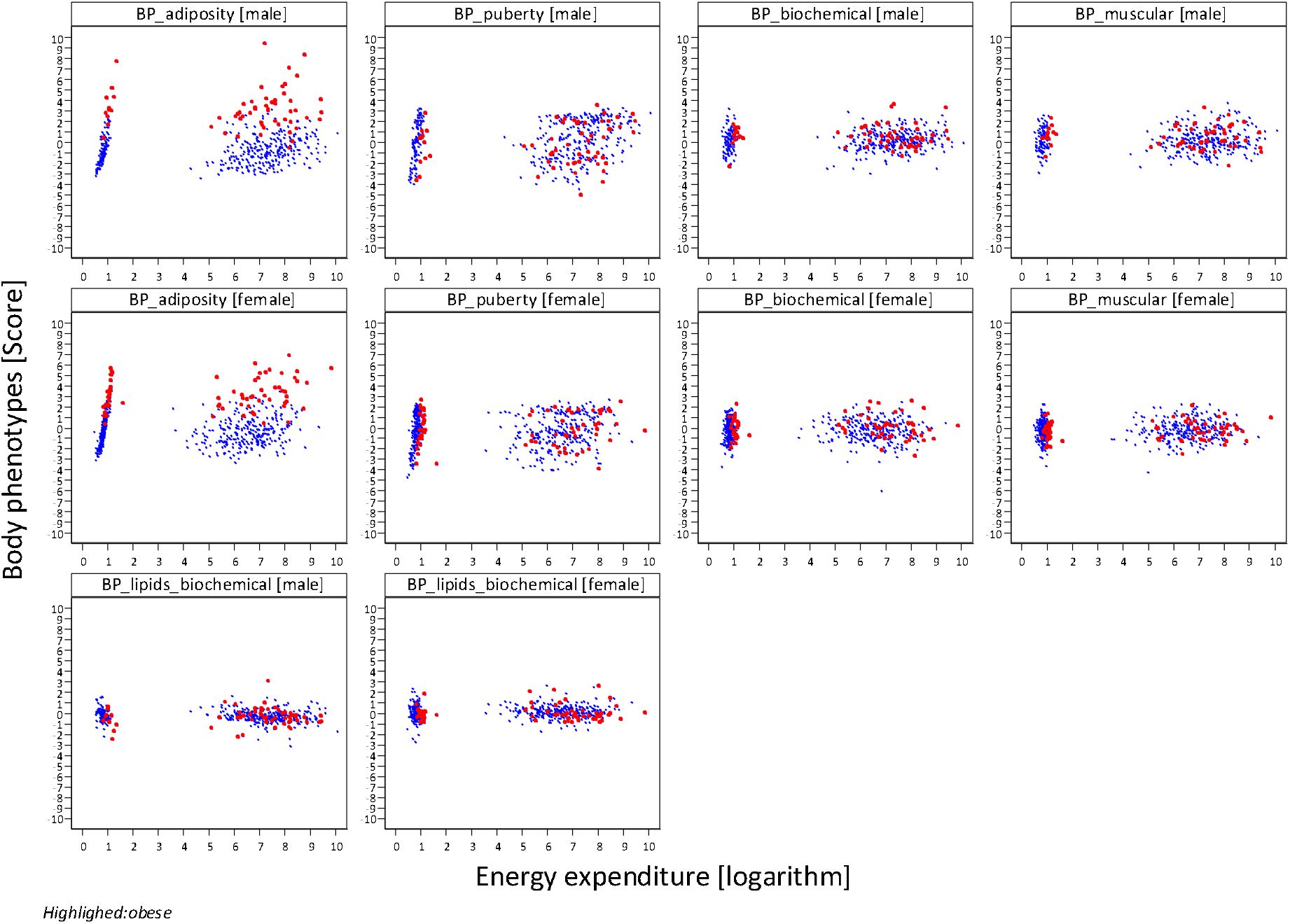
Characterization of energy expenditure and all five body phenotypes (BP) according to sex and obesity classification (red points). Piracicaba, São Paulo

Five DPs were defined for food groups. The first DP (ultra-processed foods) was composed mainly of ultra-processed foods; the second (fresh foods) with fresh foods and culinary preparations; the third (bread_rice_beans) was made up mostly of rice and beans (a typical Brazilian dish) and bread with butter or margarine (typical foods for breakfast); the fourth (culinary_preparation) consisted of culinary preparations such as cakes, rice, beans and potatoes, and corn; and the fifth (cakes_rice_beans) included rice and beans, and cakes (Table 3).

**Table 3.** Description of dietary patterns (DP) of adolescents. Piracicaba-SP, Brazil, 2013-2014.

| Food groups | Eigenvalues |  |  |  |  |
| --- | --- | --- | --- | --- | --- |
|  | DP_ultraprocessed | DP_fresh_foods | DP_bread_rice_beans | DP_culinary_preparation | DP_cake_rice_beans |
| Fruits | 0.2026 | 0.3858 |  | -0.2922 |  |
| Vegetables |  | 0.5671 |  |  |  |
| Meat/poultry/fish/eggs | 0.2695 |  |  | -0.3262 |  |
| Processed meat/sausage | 0.2631 |  |  | -0.3946 |  |
| Snacks |  | -0.3213 |  |  |  |
| Fast foods | 0.2363 | -0.2564 |  |  |  |
| Breads |  |  | 0.4429 |  |  |
| Fried tubers | 0.2249 |  |  |  |  |
| Dairy foods | 0.2381 |  |  | 0.2745 | -0.5600 |
| Fatty foods | 0.2176 |  | 0.3655 |  |  |
| Sugary drinks | 0.2383 |  |  | -0.4395 |  |
| Sugar/pastries | 0.2016 | -0.2837 | 0.2908 |  | -0.4547 |
| Ice-cream | 0.2303 |  | -0.2580 |  |  |
| Cookies | 0.2419 |  |  |  |  |
| Bakery foods | 0.2801 |  | -0.2149 |  |  |
| Cakes |  |  |  | 0.3711 | 0.2785 |
| Side dishes | 0.3443 | 0.2220 |  |  |  |
| Rice and beans |  |  | 0.4314 | 0.2335 | 0.4669 |
| Boiled potato and corn | 0.2590 | 0.3157 | -0.2532 | 0.3137 |  |
| **KMO** |  |  | **0.8579** |  |  |

We verified the association between sociodemographic variables, physical activity and DPs, and BMI and BP (Table 4). Physical activity was positive associated with BMI, BP__adiposity_, and BP__puberty_. Female sex was positively associated with BP__adiposity_ and BP__lipids_biochemical_, while male sex was positively associated with BP__puberty_, BP__gly_biochemical_ and BP__muscule_. We observed a negative association between DP__ultraprocessed_ and BMI, and a positive one for DP__fresh_foods_. Three DPs were associated with BP__adiposity_; DP__fresh_foods_ was positively associated with BP__adiposity_; DP__ultraprocessed_ and DP__culinary_preparation_ were negatively associated with BP__adiposity_; and BP__biochemical_ was negatively associated with DP__fresh_foods_.

**Table 4.** Association of body mass index, body phenotypes and dietary patterns (DP), physical activity and sociodemographic variables. Piracicaba, São Paulo, Brazil, 2013-2014.

| Variables | Outcomes |  |  |  |  |  |
| --- | --- | --- | --- | --- | --- | --- |
|  | BMI<br>(score) | BP<sub>_adiposity</sub> | BP<sub>_puberty</sub> | BP<sub>_biochemical</sub> | BP<sub>_muscular</sub> | BP<sub>_lipids_biochemical</sub> |
| **DP<sub>_ultraprocessed</sub>** | **-0.08**<br>(-0.11;-0.05) | **-0.08**<br>(-0.13;-0.03) | -0.01<br>(-0.02;0.04) | 0.02<br>(-0.01;0.04) | 0.01<br>(-0.02;0.04) | 0.02<br>(-0.01;0.05) |
| **DP<sub>_fresh_foods</sub>** | **0.09**<br>(0.04;0.14) | **0.09**<br>(0.003;0.17) | 0.009<br>(-0.045;0.062) | **-0.05**<br>(-0.10;-0.004) | 0.01<br>(-0.04;0.06) | -0.04<br>(-0.09;0.01) |
| **DP<sub>_bread_rice_beans</sub>** | -0.06<br>(-0.13;-0.00) | -0.06<br>(-0.16;0.03) | 0.078<br>(0.017;0.139) | 0.04<br>(-0.02;0.09) | 0.02<br>(-0.03;0.07) | -0.01<br>(-0.07;0.04) |
| **DP<sub>_culinary_preparation</sub>** | -0.06<br>(-0.13;0.01) | **-0.15**<br>(-0.24;-0.04) | -0.04<br>(-0.11;0.03) | -0.03<br>(-0.09;0.03) | -0.02<br>(-0.08;0.04) | -0.002<br>(-0.06;0.06) |
| **DP<sub>_cake_rice_beans</sub>** | -0.02<br>(-0.08;0.05) | 0.01<br>(-0.10;0.12) | 0.02<br>(-0.05;0.08) | 0.03<br>(-0.04;0.09) | 0.01<br>(-0.06;0.07) | -0.03<br>(-0.09;0.04) |
| Physical activity | **0.24**<br>(0.16;0.32) | **0.29**<br>(0.16;0.41) | **0.11**<br>(0.03;0.19) | 0.03<br>(-0.04;0.10) | 0.03<br>(-0.05;0.10) | -0.06<br>(-0.14;0.01) |
| **Age (years)** |  |  |  |  |  |  |
| 10 to 14 | 0.00 (Ref) | 0.00 (Ref) | 0.00 (Ref) | 0.00 (Ref) | 0.00 (Ref) | 0.00 (Ref) |
| 15 to 19 | -0.20<br>(-0.34;-0.05) | **0.66**<br>(0.43;0.89) | **2.72**<br>(2.57;2.86) | -0.17<br>(-0.30;-0.04) | 0.08<br>(-0.05;0.21) | -0.07<br>(-0.14;0.12) |
| **Sex** |  |  |  |  |  |  |
| Male | 0.00 (Ref) | 0.00 (Ref) | 0.00 (Ref) | 0.00 (Ref) | 0.00 (Ref) | 0.00 (Ref) |
| Female | 0.09<br>(-0.06;0.24) | **0.41**<br>(0.18;0.63) | **-0.29**<br>(-0.43;-0.14) | **-0.46**<br>(-0.59;-0.33) | **-0.50**<br>(-0.63;-0.37) | **0.37**<br>(0.24;0.50) |
| Wealth (score) | -0.01<br>(-0.05;0.03) | 0.02<br>(-0.04;0.08) | 0.003<br>(-0.04;0.04) | **-0.05**<br>(-0.08;-0.01) | **0.05**<br>(0.01;0.08) | 0.02<br>(-0.01;0.06) |

## Discussion

In this study, we applied multivariate analysis to estimate the outcomes (BPs) and the main predictors (DPs). We highlighted the first two BPs that express adiposity and body volume, and linear growth (chronological axis of adolescence), respectively. We identified five DPs. The first of them was composed mainly of ultra-processed foods and in three of them rice and beans were identified. DP__ultraprocessed_ and DP__fresh_foods_ were negatively associated with BP__adiposity,_ and DP__culinary_preparation_ was positively associated with BP__adiposity_ for girls, and adolescents aged 15 to 19 years. DP__fresh_foods_ was negatively associated with BP__biochemical_ for girls and wealth score.

The multivariate analysis applied in our study may be considered an innovative approach to assess nutritional status. This proposal based on the multidimensionality of parameters of nutritional status (BPs) allowed us to explore multiple interactions among anthropometric, body composition, and biochemical variables.^9,10^ Also, a positive aspect of multivariate analysis is the absence of cut-off point for anthropometric measurements, body composition, and biochemical data. These biological measurements were used in our analysis without assumptions, and BP analysis is reproducible in other adolescent populations.^25^

The perspective of analysis to investigate the relationship between DPs and nutritional status indicators presented in our study is still recent and unprecedented in the type of proposal presented here. Although in recent years the use of DPs has been increasingly applied to examine the association between diet and health outcomes, linking specific DPs to chronic diseases, including obesity and related phenotypes such as body composition and cardiometabolic markers^26-30^ is innovative, most studies still prioritize the use of BMI or obesity phenotypes (weight, waist circumference, and lipid levels) in isolation to assess nutritional status.

We observed a negative association between DP__fresh_food_ and BP__biochemical_. Shang et al.^31^ (2012), in a study including 5267 children and adolescents (6 to 13 years old), found a positive association between the western dietary pattern and higher levels of triglycerides and glucose. However, mean triglycerides and glucose values are similar between healthy and western dietary patterns. There was no association between DPs and the presence of hypertriglyceridemia or elevated glucose. In another study, higher scores for healthy eating patterns were associated with lower glucose levels.^32^

Several studies have already investigated the relationship between eating behaviors and nutritional status indicators in both young people,^33-37^ and adults.^38-40^ The results of these studies showed in general that unhealthy eating practices, characterized by the presence of ultra-processed foods, high in free sugar, saturated and trans fat, depleted in protein, fiber, and most micronutrients; these refined products increase the risk of elevated body weight. In contrast, the presence of fruits, vegetables, whole grains, and nuts are protective against gains in body fat. It is important to note that the relationship between DPs and the indicators of nutritional status is better established among adults. In children and adolescents, some reviews^41,42^ have highlighted that the results are inconsistent in cross-sectional studies.

Contrary to expectations, our study identified a negative association between the DP marked by the presence of ultra-processed foods and culinary preparations, and a positive association between the DP marked by fresh food and the adiposity phenotype. DP__ultraprocessed_ was the first principal component in our study, and we observed a high percentage of adolescents that consumed ultra-processed foods identified in this DP such as processed meat, fast foods, and sugary drinks. In a study conducted with adolescents in the city of São Paulo, Brazil, the ‘healthy’ DP was also associated with the obesity profile.^37^ In a study carried out in the United States of America (Project EAT – Eating Among Teens) a positive association was observed between the “sweet and salty snacks pattern” and the risk of overweight / obesity in boys, and higher scores for the “fruit pattern” were positively associated with risk of overweight / obesity in younger boys (mean age = 12.9 years); for girls, these associations were the opposite. The authors hypothesized that food frequency questionnaire might not reflect all the foods consumed by adolescents included in the study and, therefore, DPs did not show a clear association with weight status. Another hypothesis raised was that food consumption was not the main determinant of weight status in adolescents.^43^

In a systematic review and meta-analysis study, the authors reported higher scores for unhealthy DPs, and higher values for cardiometabolic risk factors (body weight, waist circumference, lipid profile, and blood glucose). However, healthy patterns were also associated with higher values of BMI and waist circumference.^43^ The authors highlighted publication bias in their study, and due to an unexpected or implausible association between healthy patterns and higher cardiometabolic risk, and unhealthy patterns and lower cardiometabolic risk, it may not be published.^43^ Also, the protective effect of healthy DPs in adolescents may be unclear and more studies are necessary to understand the relationship of diet and nutritional status outcomes.

Considering that the relationship between unhealthy eating behaviors and negative health outcomes is well established in the literature, a possible explanation for the inverse associations found in our study would be the occurrence of reverse causality, common in cross-sectional studies in which exposure is measured concomitantly with the outcome.

The main strength of our study is its originality. To date, to the best of our knowledge, this is possibly the first study to address both the domain of food consumption and nutritional status in a multidimensional manner, and the first to analyze the association between BPs as latent variables in the model. In addition, the food frequency questionnaire applied to assess food consumption was validated for the study population.^19,20^ We also highlighted adjusted regression analysis by energy expenditure, sex, age and wealth status. A systematic review study on DPs and cardiometabolic risk in children and adolescents discussed the importance of adjusting analysis for these factors.^44^

Among the limitations of the present study, we highlight a) the cross-sectional design that prevents the attribution of causality between variables and presents the possibility of the occurrence of reverse causality, as can be seen in this study; b) the absence of some foods in the list of the food frequency questionnaire and the presence of ultra-processed and processed foods in the same food groups (i.e., pasta and noodles, and homemade cake and industrialized cake). It is important to note that when the questionnaire was developed, there was no classification based on the extent and purpose of industrial food processing, this is a recent proposal for food classification; and c) the overestimation of physical activity by adolescents. In our questionnaire there was a list of all types of physical activity, and adolescents may have overestimated. In contrast to our study, the prevalence of physically active Brazilian adolescents (with 300 minutes or more of exercise a week) in 2012 and 2015 was low, 21% and 20.7%, respectively.^37^ Furthermore, overweight and obese adolescents could be practicing physical activity to lose body weight. Finally, we highlighted possible residual confounding effects even after adjustments for main factors (age, sex, wealth, and energy expenditure).

Little is known about the associations of DPs in relation to obesity, metabolic risk, or both among young people in emerging economies such as Brazil, who may be at greater risk for the development of obesity and obesity-related diseases due to rapid changes in the food environment and ease of access to unhealthy processed foods. Thus, the multidimensional analysis of the parameters of nutritional status and its relationship with DPs should be further explored for a better understanding of this association, since it is an innovative approach.

## Data Availability

I confirm the availability of all data referred to this manuscript

